# Drivers of bias in diagnostic test accuracy estimates when using expert panels as a reference standard

**DOI:** 10.1101/2023.07.26.23293187

**Authors:** BE Kellerhuis, K Jenniskens, E Schuit, L Hooft, KGM Moons, JB Reitsma

## Abstract

**Objectives:** To assess the impact of study and expert panel characteristics on index test diagnostic accuracy estimates.

**Study Design and Setting:** Simulations were performed in which an expert panel was used as reference standard to estimate the sensitivity and specificity of an index diagnostic test. Diagnostic accuracy was determined by combining probability estimates of target condition presence, as provided by experts using four component reference tests, through a predefined threshold. Study and panel characteristics were varied in several scenarios: target condition prevalence (20%, 40%, 50%), accuracy of component reference tests (70%, 80%, mixed), expert panel size (2, 3, 10), study population size (360, 1000), and random or systematic differences between expert’s probability estimates. Bias in accuracy estimates across all possible true index test values was quantified for all scenarios. The total bias in each scenario was quantified using the mean squared error (MSE).

*Results:* When estimating an index test with 80% sensitivity and 70% specificity, bias in these estimates was hardly affected by the study population size or the number of experts. When one expert was systematically biased, bias in sensitivity and specificity estimates increased, but this effect lessened when the number of experts in the panel was higher. Prevalence had a large effect on bias, scenarios with a prevalence of 0.5 estimated sensitivity between 63.3% and 76.7% and specificity between 56.1% and 68.7%, whereas scenarios with a prevalence of 0.2 estimated sensitivity between 48.5% and 73.3% and specificity between 65.5% and 68.7%. Random and systematic differences between experts also increased bias, with estimated sensitivity between 48.6% and 77.4% and specificity between 59.1% and 69.1% as opposed to scenarios without random or systematic differences, which estimated sensitivity between 58.0% and 77.4% and specificity between 56.1% and 69.1%. More accurate component reference tests also reduced bias. Scenarios with four component tests of 80% sensitivity and specificity estimated index test sensitivity between 60.1% and 77.4% and specificity between 62.9% and 69.1%, whereas scenarios with four component tests of 70% sensitivity and specificity estimated index test sensitivity between 48.5% and 73.4% and specificity between 56.1% and 67.0%.

*Conclusion:* Bias in accuracy estimates when using an expert panel will increase if the component reference tests (combined) are less accurate. Prevalence, the true value of the index test accuracy, and random or systematic differences between experts can also impact the amount of bias, but the amount and even direction will vary between scenarios.

## Introduction

Diagnostic accuracy studies are concerned with evaluating the performance of a diagnostic test. The test being evaluated is known as the index test. Typically, an index test result is compared to a reference standard [1,2]. This reference standard determines whether a target condition (i.e., disease, condition, or health state of interest) is present or absent, and as such determines whether the index test result or condition classification is correctly true or false. For many target conditions the reference standard is not perfect, meaning it may misclassify an individual’s true target condition. When there is no single perfect reference standard available, it is common to use a combination of several imperfect tests to create a reference standard [3,4]. This reference standard, which may involve a fixed classification algorithm [5], a statistical (latent class) model [6], or an expert panel/consensus strategy [7], is used to determine the ‘true’ presence of the target condition. However, the composite reference standard itself is inherently imperfect due to the combination of these individual imperfect tests [8, 9].

Expert panels are groups of content experts that decide on the presence or absence of the target condition in an individual. Typically, expert panels provide a single dichotomous classification for each individual, i.e., target condition present or absent. This is done through methods such as consensus meeting or majority voting [7,10,11]. Although dichotomous classification of the target condition by experts within a panel is currently still standard practice in research, it has been shown in simulation studies to potentially introduce bias in accuracy estimates (e.g., sensitivity and specificity) of the index test when there is substantial uncertainty regarding that panel classification, i.e., when a set of test results occurs in participants with the target condition and participants without the target condition at similar rates [12].

An alternative to dichotomous classification is to ask experts to provide probabilistic estimates on whether the target condition is present in each study participant. This prevents loss of information on uncertainty through dichotomization but introduces new challenges [13]. While previous studies have shown that dichotomous reference standard classification can cause bias [12], it is still unclear how characteristics of the study (e.g., prevalence) and expert panel (e.g., number of experts) affect the validity of diagnostic accuracy estimates of an index test. Additionally, little is known about how to optimally combine estimates from different experts in an expert panel.

In this simulation study we assessed how diagnostic accuracy estimates of an index test are influenced by various study and expert panel characteristics. Specifically, we focused on scenarios where an expert panel is used as the reference standard and experts provide probability estimates on presence of the target condition.

## Methods

This simulation study was designed using the structured ADEMP approach for planning simulation studies [14].

### Estimands

Our estimands are the ‘true’ sensitivity (Se) and specificity (Sp), which reflect the diagnostic accuracy of the index test for assessing the true target condition and the ‘observed’ sensitivity and specificity based on an expert panel as reference standard.

### Data-generating mechanism

We performed a full factorial assessment of study and panel characteristics. The following input parameters were specified: number of experts in the panel, number of study participants, target condition classification threshold, prevalence of the target condition, sensitivity and specificity of the component tests, random and systematic differences between experts, and the sensitivity and specificity of the component tests. This yields a total of 1296 scenario simulations. The parameters and their values are shown in table 1.

**Table 1.** Summary of study and panel characteristics and their values as used in simulations. Prevalence indicates the prevalence of the target condition. Random and systematic differences, when included, are drawn from a beta distribution. Se = sensitivity, Sp = specificity

| Study/Panel characteristic | Values |
| --- | --- |
| Number of experts | 2, 3, 10 |
| Number of participants | 360, 1000 |
| Classification threshold | 0.2, 0.3, 0.5 |
| Prevalence | 0.2, 0.4, 0.5 |
| Sensitivity and specificity of the component tests | 4 tests, Se 0.7, 0.7, 0.7, 0.7<br>Sp 0.7, 0.7, 0.7, 0.7<br>4 tests, Se 0.8, 0.8, 0.8, 0.8<br>Sp 0.8, 0.8, 0.8, 0.8<br>4 tests, Se 0.6, 0.7, 0.8, 0.9<br>Sp 0.9, 0.8, 0.7, 0.6 |
| Systematic differences | Yes, No |
| Random differences | Yes, No |
| Overconfident expert | Yes, No |

We used a model-based data-generating mechanism consisting of five steps. Table 2 provides a description of the relevant parameters in the data-generating process.

**Table 2.**
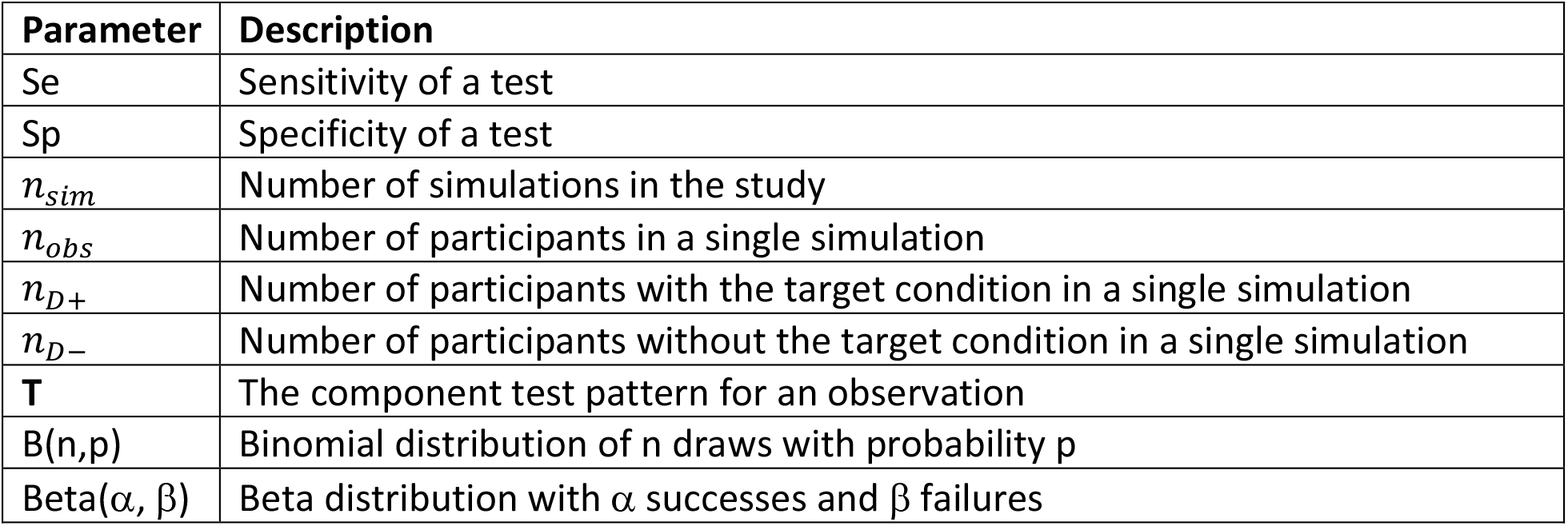
Description of variables.

| Parameter | Description |
| --- | --- |
| Se | Sensitivity of a test |
| Sp | Specificity of a test |
| $n_{sim}$ | Number of simulations in the study |
| $n_{obs}$ | Number of participants in a single simulation |
| $n_{D+}$ | Number of participants with the target condition in a single simulation |
| $n_{D-}$ | Number of participants without the target condition in a single simulation |
| **T** | The component test pattern for an observation |
| B(n,p) | Binomial distribution of n draws with probability p |
| Beta($\alpha$, $\beta$) | Beta distribution with $\alpha$ successes and $\beta$ failures |

In step 1 we simulated presence of the target condition for *n*_*obs*_ participants by drawing *n*_*obs*_ observations from a binomial distribution with success probability equal to the prevalence of the target condition (formula 1). A success (1) represented a participant with the target condition, whereas a failure (0) represents a participant without the target condition.

In step 2 the presence or absence of the target condition was used to simulate component test results. We distinguish two types of tests, component tests and an index test. In all scenarios, we will simulate 4 component tests. Component tests are used in constructing the reference standard. The reference standard we use is an expert panel, so the component tests are the information that the expert panel uses to determine the probability of an individual having the target condition. These were calculated based on the simulated true target condition status and the true sensitivity and specificity of the test, which may differ between tests. Component test results of participants with the target condition were estimated using *n*_*D*+_ draws from a binomial distribution with a success probability equal to the sensitivity of the test (formula 2.1). Results of participants without the target condition were estimated using *n*_*D*-_ draws from a binomial distribution with a success probability equal to one minus the specificity of the test (formula 2.2).

The index test is the test under review in a diagnostic accuracy study. The (binary) index was simulated in the same manner as the component tests, by drawing from a binomial distribution, in which parameter values depend on true target condition status.

In step 3 the component test results and prevalence of the target condition were used to simulate the probability estimates from the expert panel to use as the reference standard. The index test was never considered in the reference standard, only the component tests. Bayes’ Theorem (formula 3) was used to derive perfectly calibrated probabilities of target condition presence given a pattern of component test results.

In step 4, we simulated random and systematic differences between experts on the expert panel. We simulated differences by drawing from a beta distribution (formula 4). The distribution is centered around the probability of the component test results for each participant, simulating some uncertainty around that center.

We simulated random differences by drawing from the beta distribution for each participant, once per expert. The result is a new estimate with a slight uncertainty, where estimates are different for each participant. This process results in a probability estimate from each expert on each participants probability to have the target condition.

Systematic variability or differences were introduced at the level of the individual expert by calculating the probability of each possible set of component test results and then drawing from the beta distribution once per expert per set of component test results. Experts then assigned probability estimates for participants based on their component test results. This resulted in each expert having a slightly different estimate than other experts for any participant. However, unlike in the case of random differences, here participants with the same set of component test results will receive the same estimate from the expert panel.

In some scenarios step 5 was added to introduce an overconfident expert (i.e., an expert whose probability estimates skew towards 0 when below 0.5 and skew towards 1 when above 0.5). This was simulated by randomly selecting an expert in the panel who had all their probability estimates adjusted by half the distance between the estimate and either 0 or 1 (formula 5). For example, an estimate of 0.6 would be adjusted to 0.8 and an estimate of 0.2 to 0.1 when an expert is overconfident.

Figure 1 is a graphical representation of the data-generating process. For each scenario, this process is repeated 1000 times (n_sim_ = 1000).

**Figure 1.**
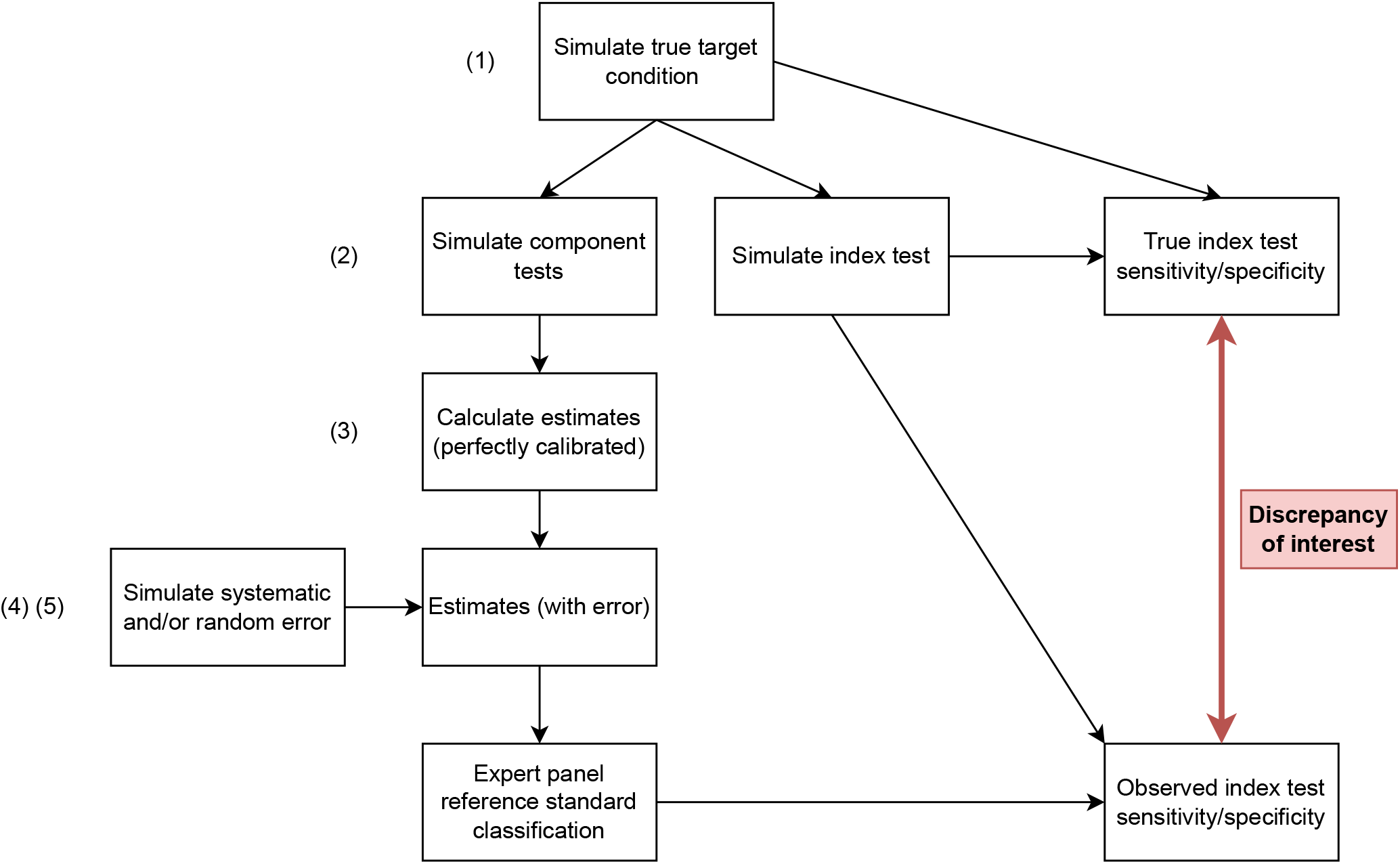
Visualization of the data-generating process. Bolded in red is the target of interest, the discrepancy between the true index test sensitivity and specificity and the observed index test sensitivity and specificity. Bracketed numbers indicate at what level each formula is applied.

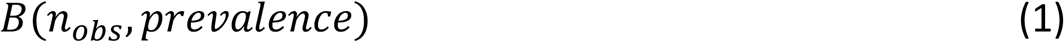

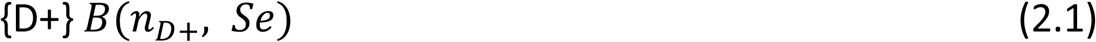

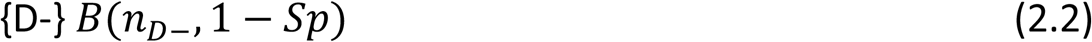

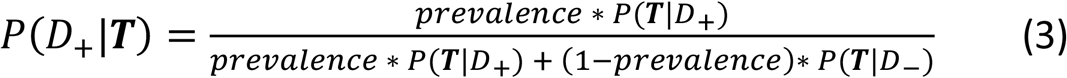

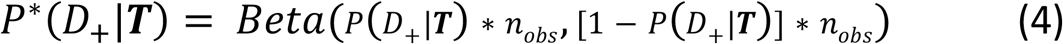

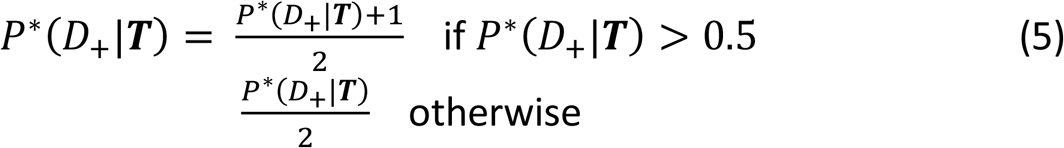

### Performance measures

We used the minimum, median, mean, and maximum of the probability estimates of target condition presence from the individual experts to come to a single panel level probabilistic estimate of target condition presence. We analysed each simulated dataset by estimating the sensitivity and specificity of the index test for the target condition as determined by applying a prespecified cut-off to the panel level probabilistic estimate.

We calculated the mean squared error (MSE) of index test sensitivity and specificity estimates calculated against the reference standard, compared to the true sensitivity and specificity calculated against the underlying target condition. In all cases, lower values indicate estimates closer to the true values.

We present our results in nested loop plots designed for clarity in reporting simulation results [15].

Additionally, we extracted a scenario with MSE near the median to illustrate the absolute bias in sensitivity and specificity of an index test across a range of true values (at increments of 0.05).

## Results

We will provide the overall impact of various scenarios on the MSE for expert panels with no random or systematic difference, only systematic differences, and both random and systematic differences. The effects of study and expert panel characteristics will be discussed. Additionally, a scenario with MSE near the median will be highlighted to illustrate the impact of using an expert panel on bias in diagnostic accuracy estimates of the index test under study.

### MSE in sensitivity estimates without random or systematic differences

Figure 2 shows the MSE of sensitivity and specificity for 1000 simulations of all scenarios without any random or systematic differences between experts (i.e., identical probabilities for each participant with the same component test pattern). The different consensus mechanisms overlap. Because there was no variability between experts, there was no difference between the different mechanisms.

**Figure 2.**
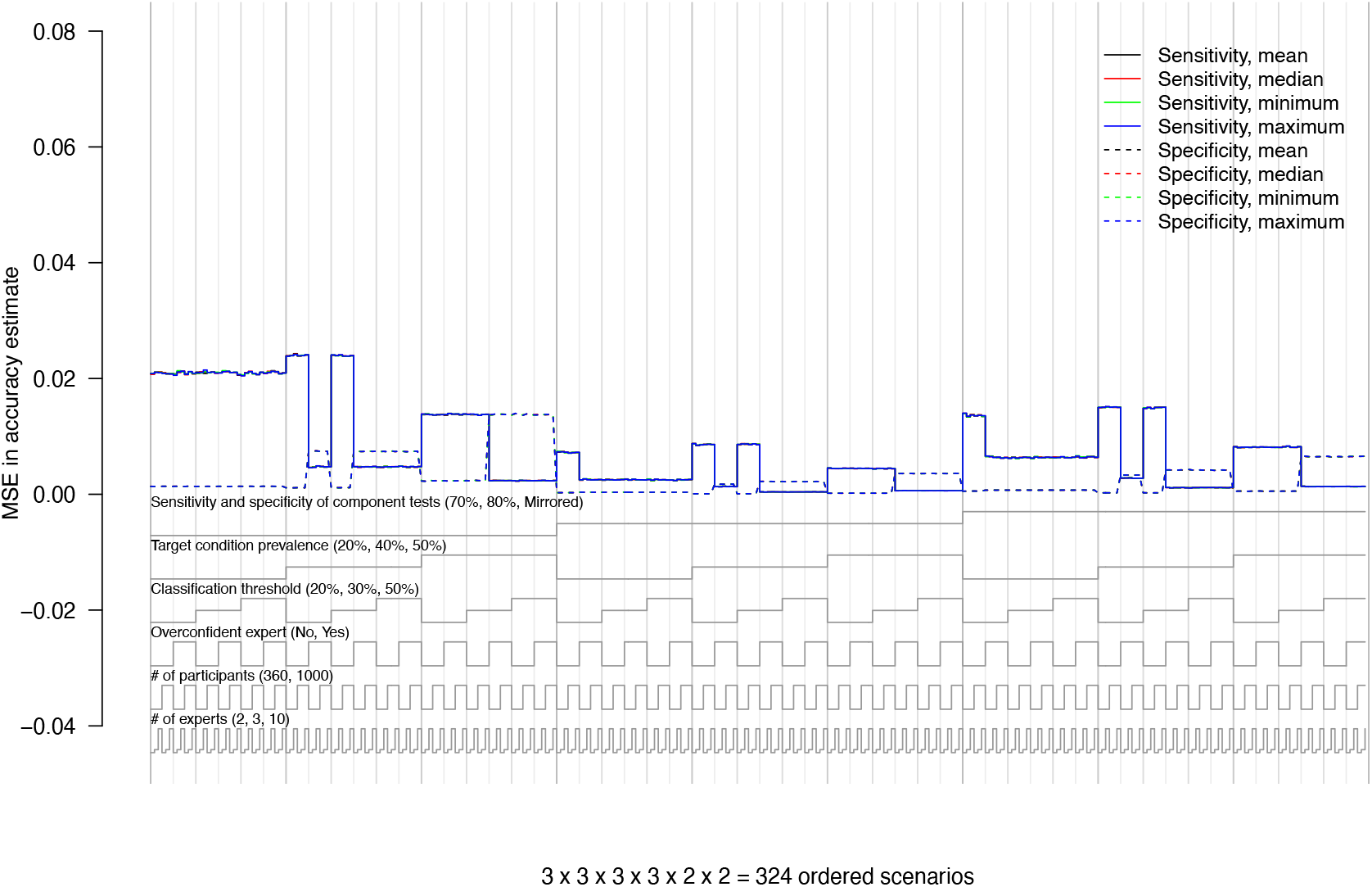
MSE in sensitivity estimates without random or systematic differences. Nested loop plot showing results of 324 expert panel scenarios without random or systematic differences between experts. Solid lines show the MSE in sensitivity and dashed lines show the MSE in specificity. Different consensus mechanisms are shown in color. At the bottom of the graph the study and panel characteristics are pictured. The different levels of the study and panel characteristics describe the scenarios corresponding to the results pictured above.

### Sensitivity and specificity of the component tests

The sensitivity and specificity of the component tests had a relatively large effect on the MSE. Scenarios with a component test sensitivity and specificity of 80% showed markedly lower MSE than scenarios where it is 70%. When the component tests are ‘mirrored’ and included two tests with low sensitivity and high specificity and two tests with high sensitivity and low specificity, MSE was in between that of 70% and 80%. There was no apparent effect of the ‘mirroring’ compared to the expected MSE for component tests of 75% sensitivity and specificity.

### Prevalence

MSE also changed with target condition prevalence. When prevalence increased, average MSE in sensitivity estimates decreased and average MSE in specificity estimates increased. The effect appeared to be relatively complex, as some combinations of scenarios increased MSE in sensitivity estimates, even though the overall direction was a decrease in MSE.

### MSE in sensitivity estimates with random and systematic differences

Figure 3 shows the MSE in accuracy estimates in scenarios with random and systematic differences between experts. The MSE was larger compared to scenarios without random or systematic differences, an increase in the highest MSE from 0.025 to 0.065.

**Figure 3.**
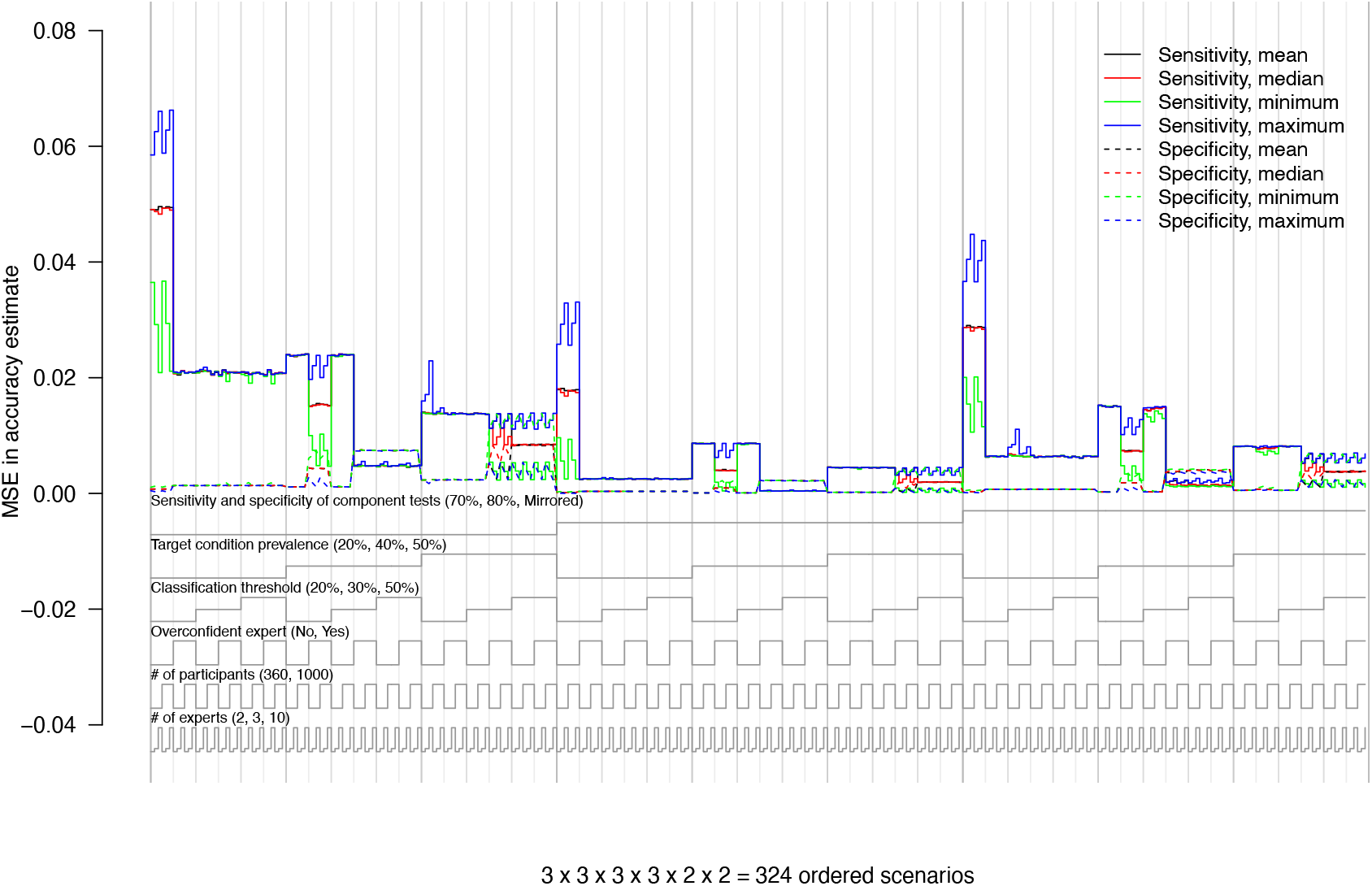
MSE in sensitivity estimates with random and systematic differences. Nested loop plot showing results of 324 expert panel scenarios with random and systematic differences between experts. Solid lines show the MSE in sensitivity and dashed lines show the MSE in specificity. Different consensus mechanisms are shown in color. At the bottom of the graph the study and panel characteristics are pictured. The different levels of the study and panel characteristics describe the scenarios corresponding to the results pictured above.

### Number of experts in the expert panel

Increasing the number of experts in the panel did not, on average, change the MSE. When the consensus mechanism was the maximum or minimum of the expert estimates, more experts lead to more extreme results. I.e., increasing the number of experts lead to higher MSE in sensitivity when the consensus mechanism was the maximum and lower MSE in sensitivity when the consensus mechanism was the minimum. The reverse was true for the MSE in specificity.

### Number of participants

Increasing the number of participants did not substantially change the MSE. There was no apparent change overall and there did not appear to be any interaction with other factors.

Figure 4 shows the observed and true sensitivity and specificity in one of the biased scenarios. This scenario had 1000 study participants, 10 experts in the panel, 40% target condition prevalence, 20% classification threshold, and sensitivity and specificity of 70% for all four component tests. In this scenario, the MSE in sensitivity was 0.015 and the MSE in specificity is 0.004. These values are also shown in figure 3.

**Figure 4.**
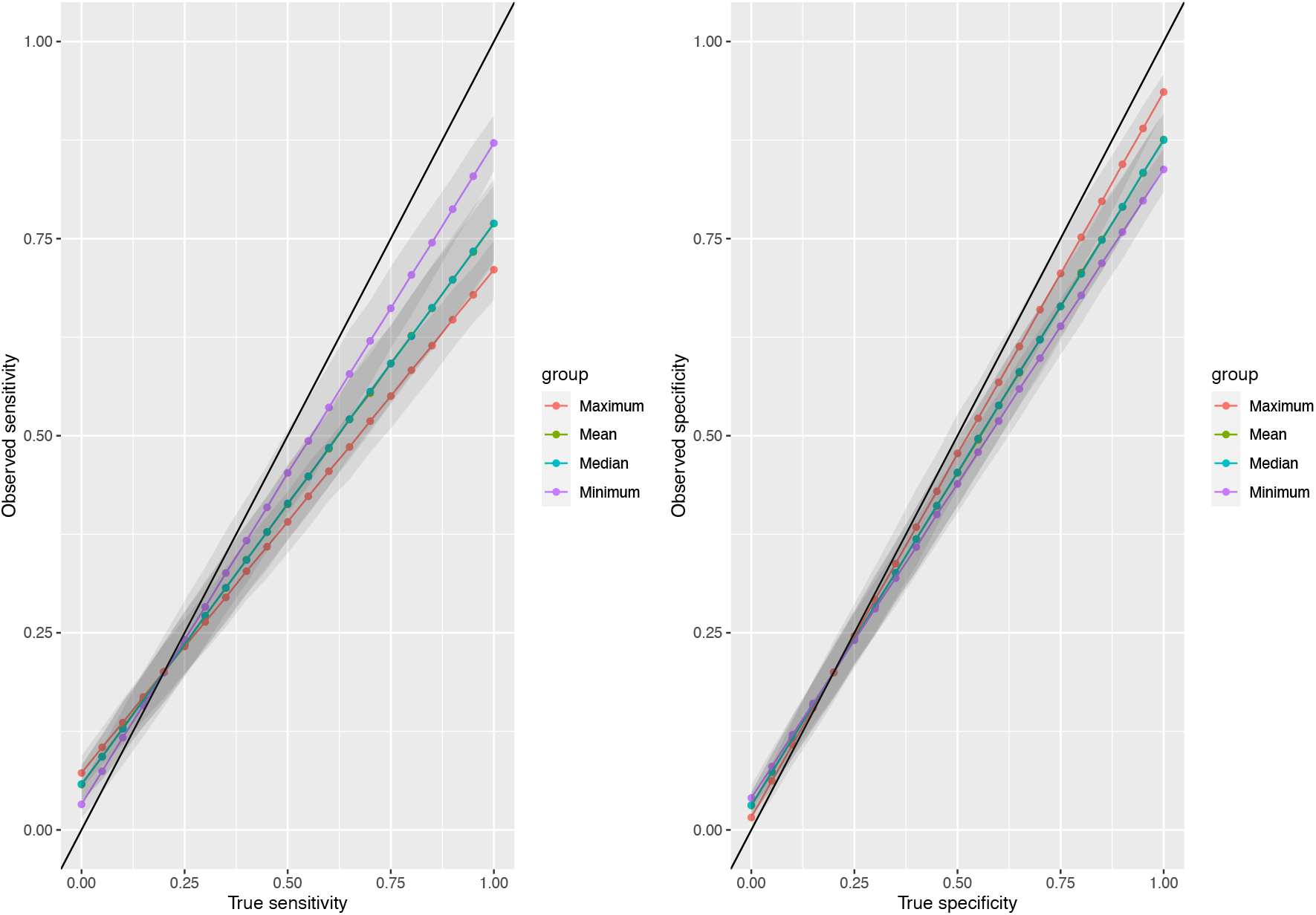
Observed and true sensitivity and specificity of a scenario with 1000 participants, 10 experts, 40% prevalence, a 20% classification threshold, 70% sensitivity and specificity in the component tests. This scenario includes random and systematic differences between experts as well as an overconfident expert. Different consensus mechanisms are presented in color with 95% intervals in grey.

The absolute differences between true and observed sensitivity are pictured. For example, given a true sensitivity of 0.75, the observed sensitivity is 0.59, hence an absolute bias of 0.16. The differences were smaller for specificity, for a true specificity of 0.75 the observed specificity is 0.70, hence an absolute bias of 0.05.

### MSE in sensitivity estimates with systematic differences

Figure 5 shows the MSE in accuracy estimates in scenarios systematic differences between experts. The MSE was larger compared to scenarios without random or systematic differences, an increase in the highest MSE from 0.025 to 0.065.

**Figure 5.**
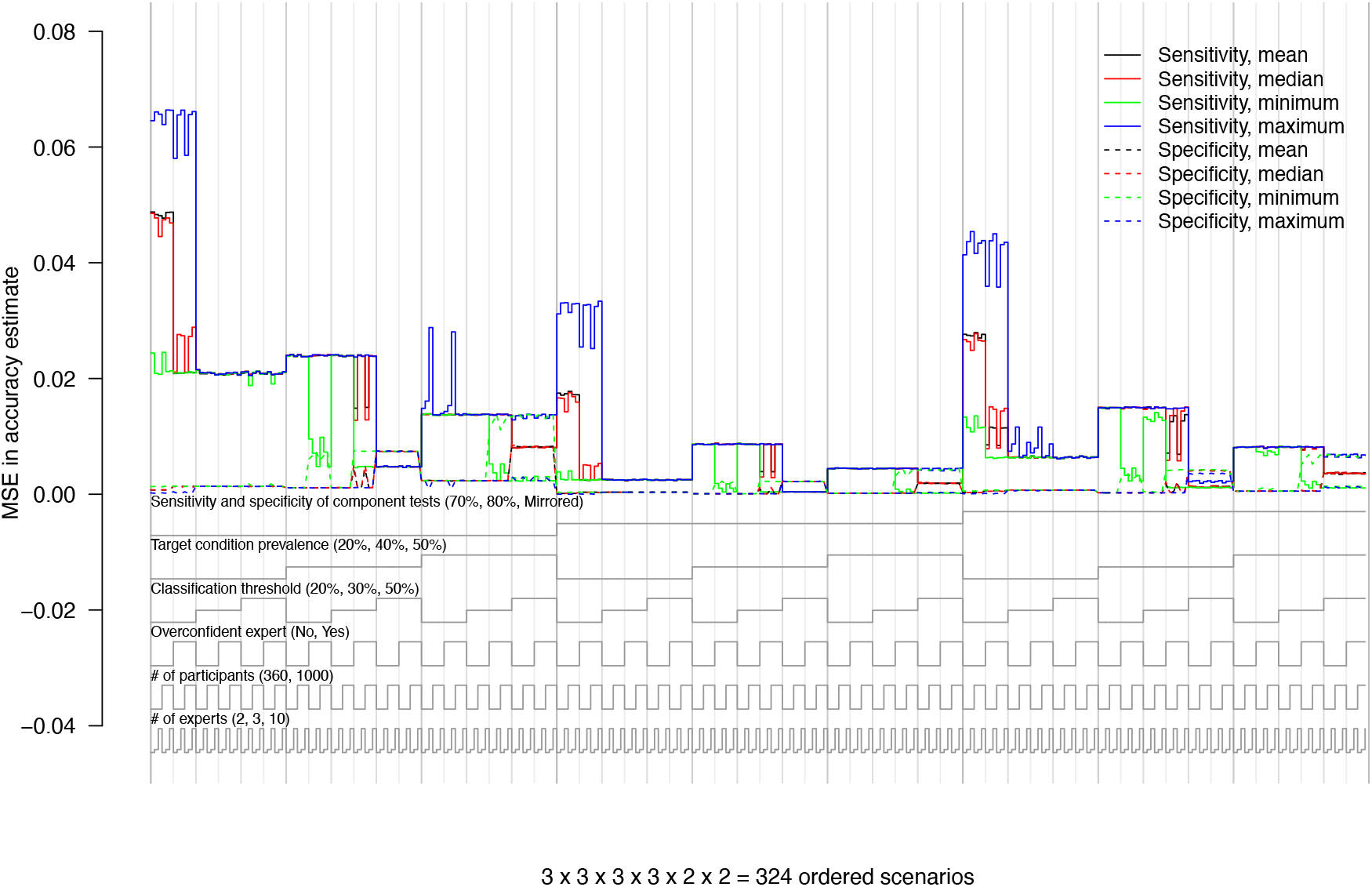
MSE in sensitivity estimates with systematic differences. Nested loop plot showing results of 324 expert panel scenarios with systematic differences between experts, but without random differences. Solid lines show the MSE in sensitivity and dashed lines show the MSE in specificity. Different consensus mechanisms are shown in color. At the bottom of the graph the study and panel characteristics are pictured. The different levels of the study and panel characteristics describe the scenarios corresponding to the results pictured above.

### Classification threshold

The choice of classification threshold had a large effect on MSE. The MSE of sensitivity was markedly higher for scenarios where the classification threshold was 20% than for scenarios where the classification threshold was 50%. Scenarios where the classification threshold was 30% sometimes had high MSE and sometimes had low MSE. Some interaction with the target condition prevalence was present. However, in no situation did a classification threshold of 30% result in a lower MSE than a classification threshold of 50%.

### Overconfident expert

In some scenarios, including an overconfident expert appeared to strongly lower MSE and in some cases an overconfident expert led to a slight increase in MSE. The effect appeared highly complex, as different combinations of factors saw highly different effects.

## Discussion

We assessed the impact of study and expert panel characteristics on diagnostic test accuracy estimates. Our results show that diagnostic test accuracy results are often biased when the reference standard is an expert panel that determines the presence of a target condition by making dichotomous classifications from probability estimates based on several component tests. Bias is reduced when the expert panel has access to more accurate component tests, but increasing the number of experts or study participants did not necessarily lead to a decrease in bias.

In most simulation scenarios the diagnostic accuracy of the index test is underestimated. Underestimation may not pose a problem if a desired minimum threshold for sensitivity or specificity is reached, for example if the test outperforms tests used in practice, but it does mean that it is not possible to infer the exact sensitivity and specificity of the test, which obscures the true value of the test. However, in some scenarios the simulation intervals also include the true value for sensitivity and/or specificity, i.e., the spread of simulated expert panels in those scenarios included overestimation and underestimation of sensitivity and specificity. So, while it is generally likely that test accuracy is underestimated when using an expert panel as reference standard, accurate estimation or overestimation is also possible. However, when true index test accuracy is high, underestimation is more likely.

Our study is in line with previously published literature. Several methods for constructing reference standards in the absence of a ‘golden standard’ have been shown to lead to potentially biased results, including composite reference standards [5], latent class models [6], and well-calibrated expert panels [12]. Despite its potential bias, expert panels are a commonly used reference standard when no gold standard reference standard is available [7].

We expand on this existing literature by exploring the impact of expert panels that are not well-calibrated. Expert panels, in practice, may suffer from random and systematic differences between experts, may include experts with different specializations, and may include experts that are more confident of their target condition assessment than is warranted. Additionally, we have performed a full-factorial analysis of all combinations of the defined input parameters. This, combined with the MSE over a series of true accuracy values of the index test, allows for inspection of bias across all possible scenarios.

When interpreting findings from our study, some limitations have to be taken into account. First, we are limited by the complex interplay in real-world expert panel meetings. While we did account for possible differences between experts, expert panels are complex systems with social dimensions. This means that they are exceedingly difficult to simulate, which meant some simplifications and approximations had to be used. A second limitation is the dichotomization of the target condition classifications (i.e., target condition present vs. absent). Expert panels are typically asked to provide target condition classifications, without reporting their own confidence in the classification. This is typically how expert panels are used in practice but is likely to result in loss of information compared to expert panels providing probability estimates for the target condition.

It is worthwhile exploring whether asking expert panels to provide probability estimates instead of dichotomized target condition classifications leads to improvements in diagnostic test accuracy estimates. To accommodate this, further research is needed on developing and validating methods to compute accuracy results based on probabilities, and to assess whether computation of accuracy estimates from probabilities, instead of classifications, leads to less biased results.

In conclusion, our study highlights the large impact study and expert panel characteristics can have on bias in diagnostic test accuracy results. Both sensitivity and specificity are typically underestimated. Surprisingly, increasing the number of experts or study participants does not lead to a reduction in this bias. However, we observed that bias can be reduced by providing the expert panel with more accurate component tests. Furthermore, we suggest that asking expert panels to provide probability estimates of target condition presence, rather than asking for a dichotomous classification, may offer additional opportunities for further bias reduction in diagnostic test accuracy estimates.

## Data Availability

All data produced in the present study are available upon request to the authors.

